# Agreement of an AI tool for joint space width measurement in radiographic knee osteoarthritis: data from the LOSEIT trial

**DOI:** 10.64898/2026.06.11.26355242

**Authors:** Suleiman Mayar, Mathias Willadsen Brejnebøl, Marius Henriksen, Robin Christensen, Phillip Hansen, Henning Bliddal, Janus Uhd Nybing, Camilla Toft Nielsen, Henrik Gudbergsen, Mikael Ploug Boesen

## Abstract

**STUDY OVERVIEW:** *Background and rationale:* Knee osteoarthritis (KOA) is a leading cause of lower limb disability worldwide, characterized by functional limitations, stiffness and pain. The incidence of KOA is especially tied to age and obesity. It is a disabling disease that often makes patients less physically active, thus increasing the risk of other diseases and mortality^1^. The clinical diagnosis of KOA is based on the symptoms and functional limitations of the joint. The diagnosis is usually supported with a radiograph (X-ray) of the weight-bearing knee. Radiographic features, such as Kellgren-Lawrence grade, are used as eligibility criteria for clinical studies while other features, such as joint space width (JSW), are used as endpoints for structural KOA progression^2,3^. While the use of these radiographic features is standard in academia, the use of JSW as a structural biomarker has received criticism. Critics point out that JSW is an indirect and projection dependent measure of cartilage deterioration which is sensitive to technical factors such as the angulation of the X-ray beam and the positioning of the knee. Small differences in these factors can alter the measured joint space and may not reflect true disease progression^4,5^. Despite limitations, minimum joint space width (mJSW) remains as one of the most widely used structural biomarkers in KOA trials and is currently one of the only structural imaging accepted in regulatory guidance as evidence of disease modification in OA drug development^3^. For JSW to be reliable and consistent in determining the advancement of KOA, the use of fixed-flexion devices is crucial to reduce the risk of unwanted narrowing or widening of the radiographic joint space width^6,7^. The LOSEIT trial, which the present study is based on, acknowledges the angulation problem and uses a standard clinical fixed-flexion device in weight-bearing PA views to get reliable JSW results^8^. Historically, a radiologist would draw on and grade radiographs of the knee-joint to extract the features. However, manual reading and annotation is time consuming with notable interobserver variance^9^. With increasing computational power and the use of deep neural networks, off-the-shelf artificial intelligence (AI) tools have become available for automatic extraction of radiograph features. Automation would free up time from radiologists and provide more consistent measurements due to the reproducible nature of the models^10^. These tools have received regulatory approval for commercial use, however, regulatory approval does not guarantee uniform or bias free performance when used on real-world data^11^. Furthermore, in a large multi-hospital chest X-ray study, Zech et al., showed that convolutional neural networks achieved worse results on data from other hospitals than on the original hospitals in which it was tested^12^. This highlights the risk of overestimating the accuracy of AI tools when only internally validated. It is therefore apparent that external validation is required when testing these AI models.

*Objectives:* The aim of this analysis is to evaluate the agreement of a commercially available AI tool for measuring JSW with the best practice radiologist annotation in the tibiofemoral joint of the knee in radiographs stabilized with a fixed-flexion device and acquired as part of a clinical trial.

*Methods:* This study is a secondary analysis of the data from the LOSEIT trial, a randomized, double-blind, placebo-controlled, single-center trial, where patients were randomized to either liraglutide or identically appearing placebo after an initial weight-loss period to investigate the effects on KOA. Radiographs of the tibiofemoral joint were acquired at enrollment (week -8) and at end-of-trial (week 52) for a total acquisition-to-acquisition time of 60 weeks^13^. The primary analysis will assess agreement between AI-derived and reference-derived change in JSW from enrolment to follow-up. Change will be calculated as follow-up minus enrolment separately for the AI tool and the reference measurement. The main measure of interest will be the change in medial minimal JSW (mmJSW), with change in lateral minimal JSW (lmJSW), medial fixed JSW (mfJSW) and lateral fixed JSW (lfJSW) as secondary measures. This study will follow an equivalence framework using the two one-sided tests (TOST) approach with a Bland-Altman analysis as the main outcome. The equivalence margin will be set at δ = 0.5 mm. Agreement consistent with equivalence will be considered established if the upper limit of the 95% confidence interval (95% CI) for the upper limit of agreement (LoA) and the lower limit of the 95% CI for the lower LoA are within the established margins. The reference JSW will be the average measurement of two independent resident radiologists. If there is a mismatch in the measurements of more than 0.40 mm between the two radiologists, the radiologists will re-annotate the case independently. If the difference remains greater than 0.40 mm, a musculoskeletal radiology consultant will review the radiograph and establish the reference JSW. The index test will be the measurements output by the AI tool.

*Populations:* Patients aged 18 to 74 with symptomatic knee osteoarthritis, radiographically confirmed KL grade 1-3, with a BMI ≥27, motivated for weight loss and in accordance with the LOSEIT trial inclusion criteria

*Further statistical details:* Sample size
Not applicable as this is a secondary analysis. Framework
This is an agreement study assessing the equivalence of a commercially available AI tool for radiographic evaluation of knee osteoarthritis with best practice radiologist measurements. Confidence intervals and P values
All 95% confidence intervals and P-values will be two-sided. Statistical software
SAS Studio and/or R version 4.2.2 (or newer).

## 2. VISUAL PRESENTATION OF TIMING OF OUTCOME MEASUREMENTS

**Figure 1.**
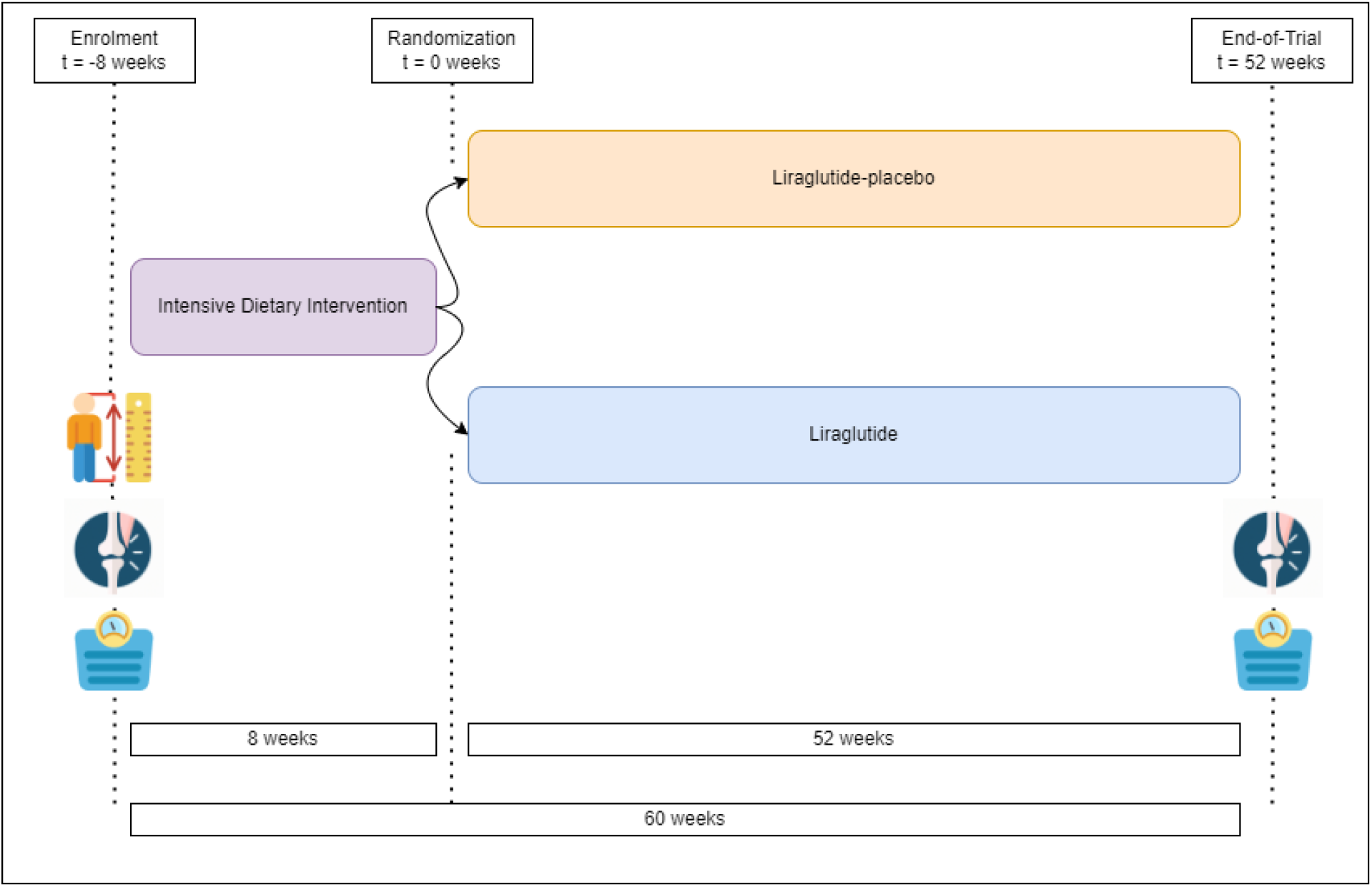
Timeline of outcome measurements in the LOSEIT trial Participants had their height and body weight measured, and underwent x-ray of the knee, at enrolment. After enrolment, they received an intensive dietary intervention for 8 weeks. If they lost more than 5% of their body weight, they were invited to be randomized to either liraglutide or placebo for 52 weeks. At the end-of-trial (60 weeks from enrolment) the participants had their body weight measured again and underwent another x-ray of the knee.

**Figure 2.**
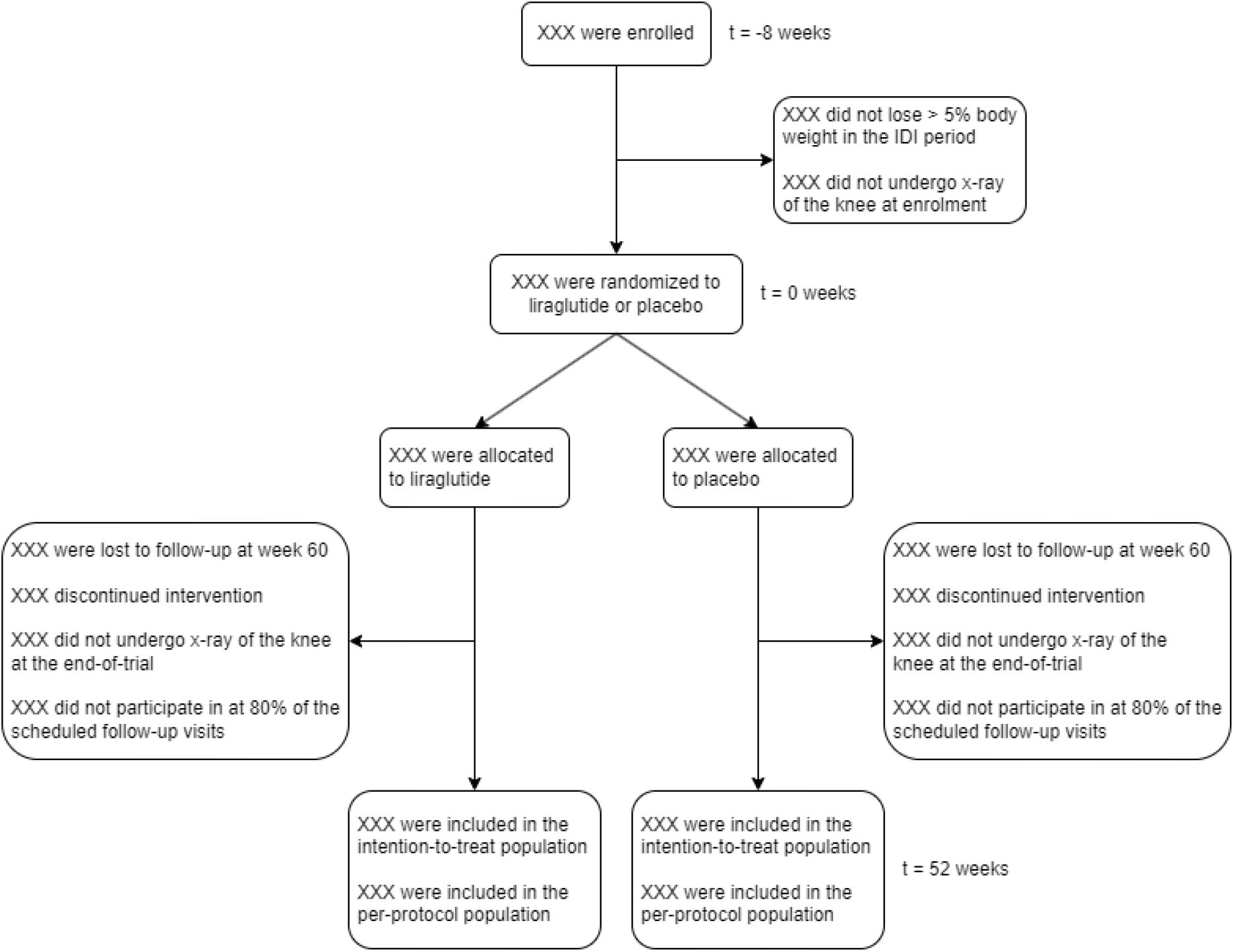
Flow diagram. Anticipated flow, illustrating potential reasons for exclusion The analysis population will be participants with measurements available for both enrolment and follow-up.

## 3. ELABORATIONS ON OUTCOMES AND DATA

### Data management

Values output by the AI tool will be compared to the reference standard.

### Measurements of reference standard

The reference standard will be established by two resident radiologists independently annotating the bony femoral and tibial surfaces of the tibiofemoral joint, and the knee coordinate system (as per Neumann and Duryea et al^14^) using a WACOM One 13” external screen with digitizer. Based on the annotations, the minimal joint space width values will be calculated, and, with assistance from the knee coordinate system, the fixed-location joint space width values will be calculated. If the difference between the radiologists is less than 0.4 mm, the average of the measurements is used. If the difference between the measurements of the two annotators is greater than 0.4 mm, the annotators will re-annotate the case independently and the evaluation is repeated. If the difference is still greater than 0.4 mm, a senior musculoskeletal radiologist will arbitrate the reference value for that measure.

#### Minimal Joint Space Width (mJSW)

Medial (mmJSW) and lateral (lmJSW) compartments.

If bone-on-bone configuration is true for the compartment, mJSW is 0. Otherwise, it is calculated as the smallest distance between femoral condyle and the tibial plateau annotations for each compartment.

#### Fixed-location Joint Space Width

Medial (mfJSW) and lateral (lfJSW) compartments will be acquired as described by Neumann and Duryea et al.

### Measurements of the index test

The AI tool RBknee-FDA will analyze the radiographs of the participants in a sequential manner. It outputs several features which can be directly read as, or derived into, the mmJSW, mfJSW, lmJSW and lfJSW. Additionally, the tool may fail to analyze a radiograph which will be noted. The tool runs locally as a containerized process.

### Data validation

All variables used in the analyses, including the derived variables, will be checked for missing values, outliers, and inconsistencies.

### Data template

Based on this SAP, the statistical analyst will develop a tailored data template illustrating the data structure required for the statistical analyses.

## 4. OUTLINE

**Table 1.** Characteristics in the LOSEIT trial population.

| Characteristics | Overall<br>(n=) |
| --- | --- |
| Age |  |
| Female sex, no (%) |  |
| Height (cm) |  |
| Body weight (kg) |  |
| Baseline BMI (kg/m2) |  |
| KOOS score |  |
| KL grade |  |

Statistical information related to Table 1: Data will be presented as means with standard deviations (SD) when normally distributed or as medians with interquartile range in case of skewed data. Dichotomous and categorical data will be presented as absolute counts and proportions.

### 4.2: Equivalence

**Figure 3.**
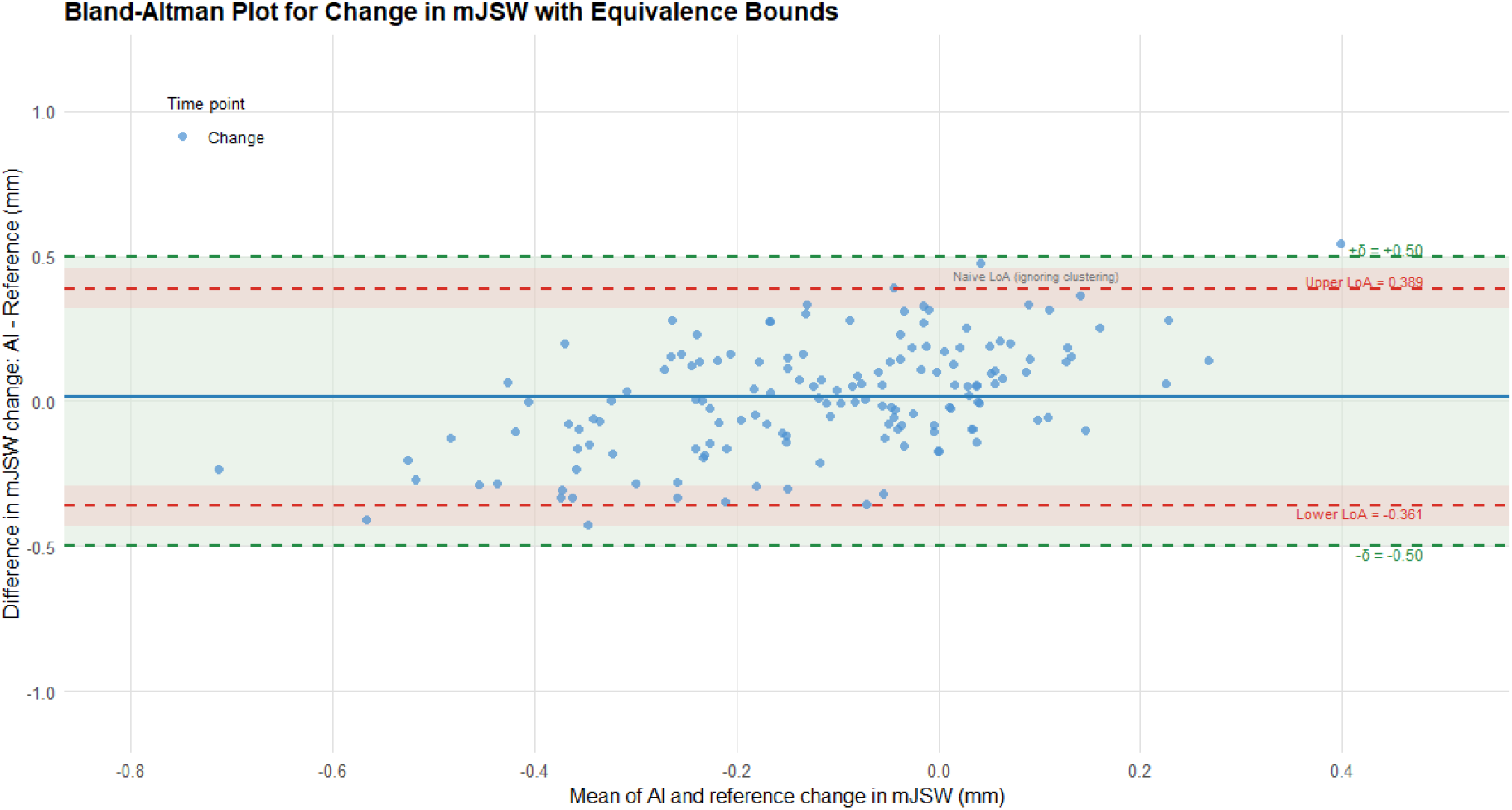
Equivalence test for agreement in JSW change between AI tool and reference (MockUp) Bland-Altman plot showing agreement between AI-derived and reference-derived change in mmJSW from enrolment to follow-up. The green area defines the predefined margins of δ = ±0.5 mm for the LoA. If the upper limit of the 95% CI of the upper LoA and the lower limit of the 95% CI of the lower LoA are both within the green area, as illustrated in this mock-up, agreement is established. If the outer limits of the 95% CI of LoA are outside the green area, agreement is not established. This test will be performed for all four joint space width measurements (medial and lateral minimal and fixed).

#### Data handling

The primary analysis population will include participants with eligible target knee radiographs and both AI and reference JSW measurements available at enrolment and follow-up. Only the target knee, as defined by the patient at enrolment, will be included. All data will be managed according to existing privacy regulations.

#### Supplementary analysis

Supplementary mixed-effects Bland-Altman analyses will be performed separately at enrolment and at follow-up for all JSW measures. Furthermore, three separate supplementary analyses will be performed.

1. Without bone-on-bone configuration: as described above except that radiographs with bone-on-bone configuration (mJSW of 0 mm) will be excluded. This is done to evaluate if the AI model performs better when not taking these radiographs into account, as the model has shown to perform worse particularly with bone-on-bone radiographs.
2. Naïve approach: as described above but without the mixed-effects component at enrolment and follow-up
3. Sensitivity analyses: as described above but with other margins (δ = 0.35 mm, δ = 0.70 mm, and δ = 0.85 mm)

##### Further statistical information related to Table 2

For Table 2, change is defined as follow-up minus enrolment for both the AI tool and the reference measurement

**Table 2.**
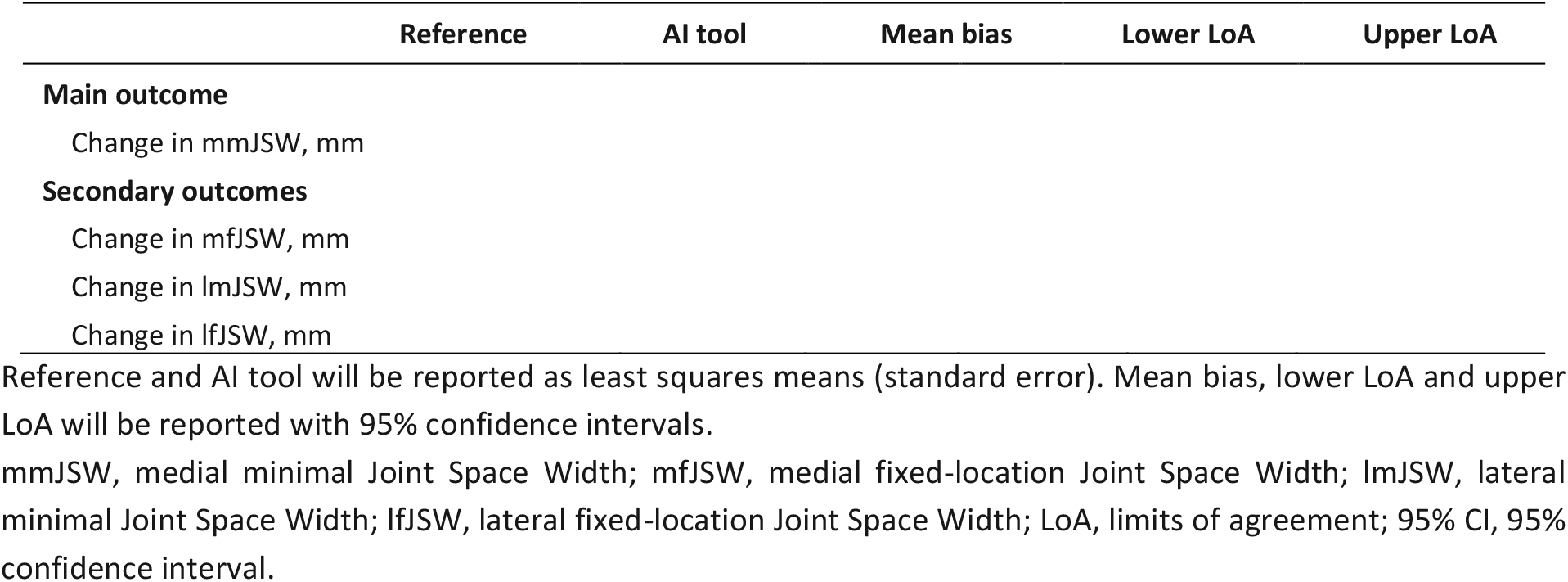
Difference in change in joint space width between the reference measurements and the AI tool.

Assumptions for the Bland-Altman models will be checked for proportional bias and homoscedasticity across measurement magnitude. The mixed-effects assumptions will be assessed by visual inspection of residual plots, including assessment of residual normality. If deviations are observed, these will be reported and considered in the interpretation.

## Data Availability

All data produced in the present study are available upon reasonable request to the authors

## SUPPLEMENTARY MATERIAL

The anticipated (predefined) supplementary material of the manuscript is illustrated below.

**Supplementary file 1. Protocol**^**8**^

**Supplementary file 2. This SAP**

**Supplementary file 3. Primary trial publication**^**15**^

**Supplementary Table 1.**
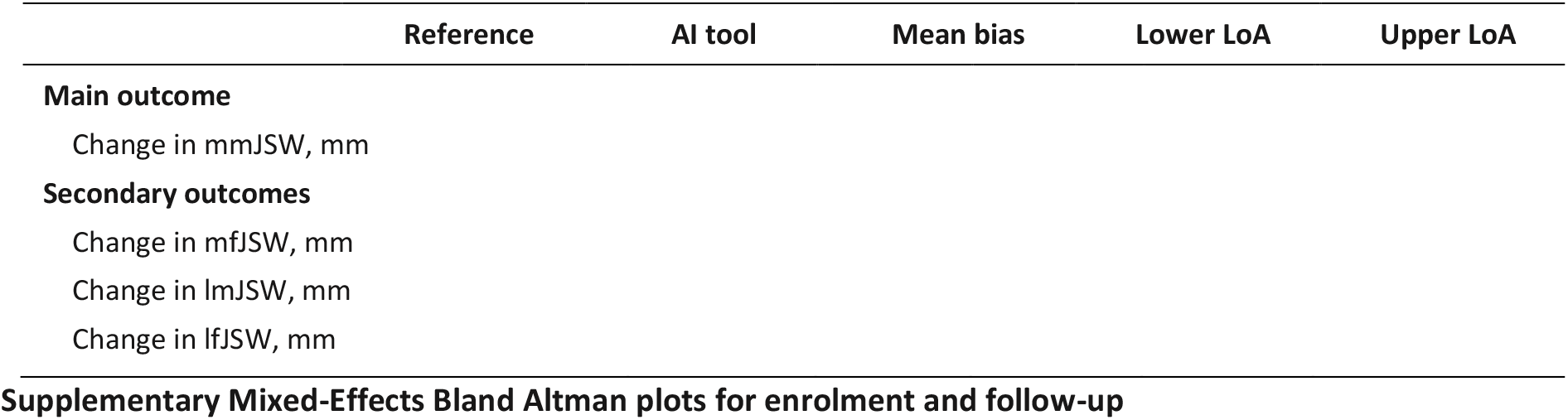
Difference in change in joint space width between the reference measurements and the AI tool when bone-on-bone radiographs are excluded.

**Figure 4.**
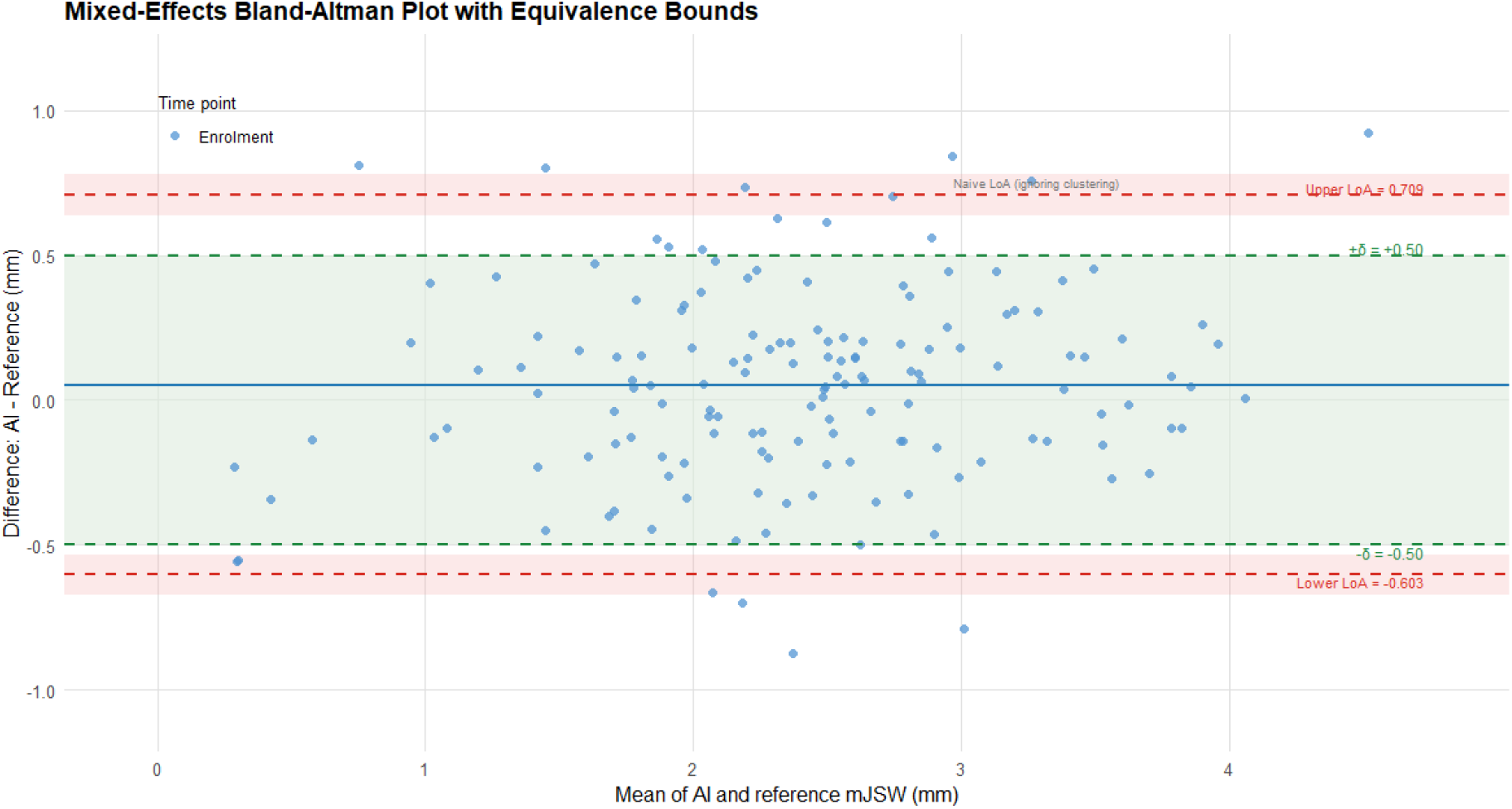
Supplementary equivalence test for JSW at enrolment between AI tool and reference (MockUp) In figure 4, the outer limits of the 95% CI of LoA are outside the green area and thus agreement is not established.

**Figure 5.**
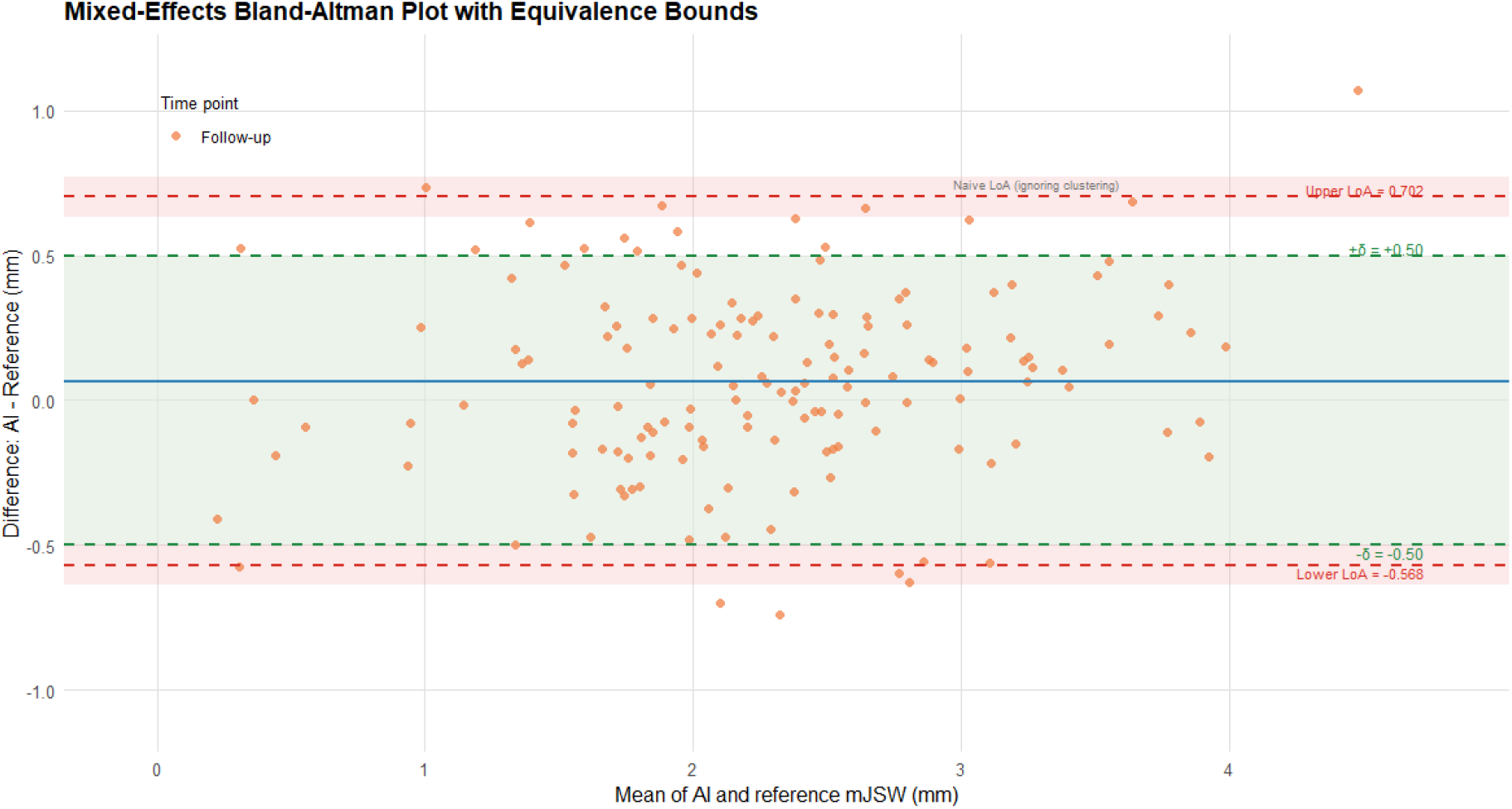
Supplementary equivalence test for JSW at follow-up between AI tool and reference (MockUp) In figure 5, the outer limits of the 95% CI of LoA are outside the green area and thus agreement is not established

## 7. SAP REPORTING GUIDELINE

This SAP has been reported according to the GRRAS guidance document: Recommended items to Address in a Reliability and Agreement Study:

| Section/Item | Index | Description | Location in this SAP |
| --- | --- | --- | --- |
| **Title and Abstract** |  |  |  |
| Title/abstract statement | 1 | State in the title or abstract that interrater/intrarater reliability or agreement was evaluated. | *Title / 1. Study overview* |
| **Introduction** |  |  |  |
| Measurement device | 2 | Clearly name and describe the diagnostic or measurement device being studied. | *1. Study overview / 3. Elaborations on outcomes and data* |
| Subject population | 3 | Specify the subject population of interest. | *1. Study overview* |
| Rater population | 4 | Specify the rater population of interest, if applicable. | *1. Study overview / 3. Elaborations on outcomes and data* |
| Existing evidence and rationale | 5 | Summarize what is already known about reliability/agreement and explain why this study is needed. | *1. Study overview* |
| **Methods** |  |  |  |
| Sample size | 6 | Explain how the sample size was determined, including planned numbers of raters, subjects/objects, and replicate observations. | *1. Study overview / 4.2 Equivalence* |
| Sampling method | 7 | Describe the sampling method. | *1. Study overview / 4.2 Equivalence* |
| Measurement/rating process | 8 | Describe the measurement/rating process, including timing between repeated assessments, access to clinical information, and blinding. | *2. Visual presentation of timing of outcome measurements / 3. Elaborations on outcomes and data* |
| Independent ratings | 9 | State whether measurements/ratings were conducted independently. | *1. Study overview / 3. Elaborations on outcomes and data* |
| Statistical analysis | 10 | Describe the statistical analysis. | *1. Study overview / 4.2 Equivalence* |
| **Results** |  |  |  |
| Included sample | 11 | Report the actual numbers of raters and subjects/objects included, and the number of replicate observations performed. | *Figure 2 / 4. Outline* |
| Sample characteristics | 12 | Describe key sample characteristics of raters and subjects/objects, such as training and experience. | *1. Study overview / 3. Elaborations on outcomes and data / 4. Outline* |
| Estimates and uncertainty | 13 | Report reliability/agreement estimates together with measures of statistical uncertainty. | *4.2 Equivalence* |
| **Discussion** |  |  |  |
| Practical relevance | 14 | Discuss the practical relevance of the findings. | N/A in SAP |
| **Auxiliary Material** |  |  |  |
| Detailed results | 15 | Provide detailed results where possible, for example in supplementary or online material. | N/A in SAP |

## Notes

### Competing Interest Statement

Mikael P. Boesen is a shareholder of Radiobotics APS.
Marius Henriksen reports a relationship with Thausne, Contura International, and Osteoarthritis and Cartilage Journal.
Henning Bliddal reports a relationship with Inc and Contura International.

### Clinical Trial

NCT02905864

### Clinical Protocols

https://pmc.ncbi.nlm.nih.gov/articles/PMC6501972/

### Author Declarations

The primary trial was approved by the regional ethics committee in the Capital Region of Denmark (H-16019969), the Danish Medicines Agency and the Danish Data Protection Agency.

### Funding

- Novo Nordisk (Denmark)
- Cambridge Weight Plan (United Kingdom)
- Oak Foundation: OCAY-13-309

